# A framework for a brain-derived nosology of psychiatric disorders

**DOI:** 10.1101/2024.05.07.24306980

**Authors:** Tristram A. Lett, Nilakshi Vaidya, Tianye Jia, Elli Polemiti, Tobias Banaschewski, Arun L.W. Bokde, Herta Flor, Antoine Grigis, Hugh Garavan, Penny Gowland, Andreas Heinz, Rüdiger Brüh, Jean-Luc Martinot, Marie-Laure Paillère Martinot, Eric Artiges, Frauke Nees, Dimitri Papadopoulos Orfanos, Herve Lemaitre, Tomáš Paus, Luise Poustka, Argyris Stringaris, Lea Waller, Zuo Zhang, Lauren Robinson, Jeanne Winterer, Yuning Zhang, Sinead King, Michael N. Smolka, Robert Whelan, Ulrike Schmidt, Julia Sinclair, Henrik Walter, Jianfeng Feng, Trevor W. Robbins, Sylvane Desrivières, Andre Marquand, Gunter Schumann, IMAGEN Consortium, environMENTAL Consortium

## Abstract

Current psychiatric diagnoses are not defined by neurobiological measures which hinders the development of therapies targeting mechanisms underlying mental illness ^1,2^. Research confined to diagnostic boundaries yields heterogeneous biological results, whereas transdiagnostic studies often investigate individual symptoms in isolation. There is currently no paradigm available to comprehensively investigate the relationship between different clinical symptoms, individual disorders, and the underlying neurobiological mechanisms. Here, we propose a framework that groups clinical symptoms derived from ICD-10/DSM-V according to shared brain mechanisms defined by brain structure, function, and connectivity. The reassembly of existing ICD-10/DSM-5 symptoms reveal six cross-diagnostic psychopathology scores related to mania symptoms, depressive symptoms, anxiety symptoms, stress symptoms, eating pathology, and fear symptoms. They were consistently associated with multimodal neuroimaging components in the training sample of young adults aged 23, the independent test sample aged 23, participants aged 14 and 19 years, and in psychiatric patients. The identification of symptom groups of mental illness robustly defined by precisely characterized brain mechanisms enables the development of a psychiatric nosology based upon quantifiable neurobiological measures. As the identified symptom groups align well with existing diagnostic categories, our framework is directly applicable to clinical research and patient care.

## Main

There has been a growing imperative within psychiatric neuroscience to uncover the biological mechanisms underlying mental health and disease to develop more effective treatments ^3,4^. A major challenge lies in the classification of psychiatric disorders since their categorization does not follow biological mechanisms. Biological links distinguishing diagnostic criteria, including brain structure ^5,6^, function ^7^, and connectivity ^8^, are limited, pointing to shared neurobiological substrates across mental illnesses. Dysfunctions within one mechanism might affect the clinical presentation of more than one diagnosis, giving rise to comorbidity^9,10^. A potential solution to this challenge is to rearrange existing clinical measures of psychopathology so they optimize the link between symptoms and biology - an approach that may lead to the discovery of novel biomarkers and targets for treatment development.

This need is perhaps most apparent by the efforts of biology-driven initiatives, such as the creation of the National Institute of Mental Health’s Research Domain Criteria (RDoC) framework ^3,4^. While RDoC offers an innovative framework, its focus is on basic biological processes linking animal and human research. In doing so, RDoC diverges from current practices used by clinicians, and arguably, the RDoC constructs may be insufficient to address the totality of mental disease. Alternatively, the Hierarchical Taxonomy Of Psychopathology (HiTOP) maintains the complex clinical characterization with focus on clinical spectrum and hierarchy ^11^. HiTOP constructs are not driven by the biology that underlies psychiatric liability. A unifying framework that takes into account both the complex biological variation and the clinical variation concurrently to characterize nosology is needed.

To address this need, we employed a data-driven strategy enabling us to integrate information from multiple domains, including clinical symptoms, brain structure (such as white matter fractional anisotropy, cortical thickness, and surface area), as well as intrinsic (resting state fMRI) and extrinsic (task fMRI) features of brain function. Our analysis aims at proposing a novel, biology-driven framework for psychiatric nosology that harnesses current clinical assessments and quantifiable neurobiological measures, such as comprehensive functional and structural neuroimaging data.

### Sparse generalized canonical correlation analysis (SGCCA) model optimization

To establish an optimized SGCCA model (Figure 1), we aimed to reduce the number of collinear variables in our data views while maximizing the variance explained. The optimal L_1_ sparsity for all data views was lambda = 0.3 after 1000 permutations at each of the 10 steps (z-statistic = 12.6, Figure 2a). We selected ten components as the point in which the full model variance explains cumulative average variance explained (AVE) levels off at 40.4% (Figure 2b). The stability selection was performed by randomly selecting 50% of the training data without replacement 10000 times and retaining the clinical items, brain regions, and resting-state brain mode connectivity variables that appeared in 90% of the subsampled SGCCA models (Figure 2cd). The final model (selected variables, ten components, and λ_1_=1.0) explained 52.7% of the variance among all data views (Figure 2e).

**Figure 1.**
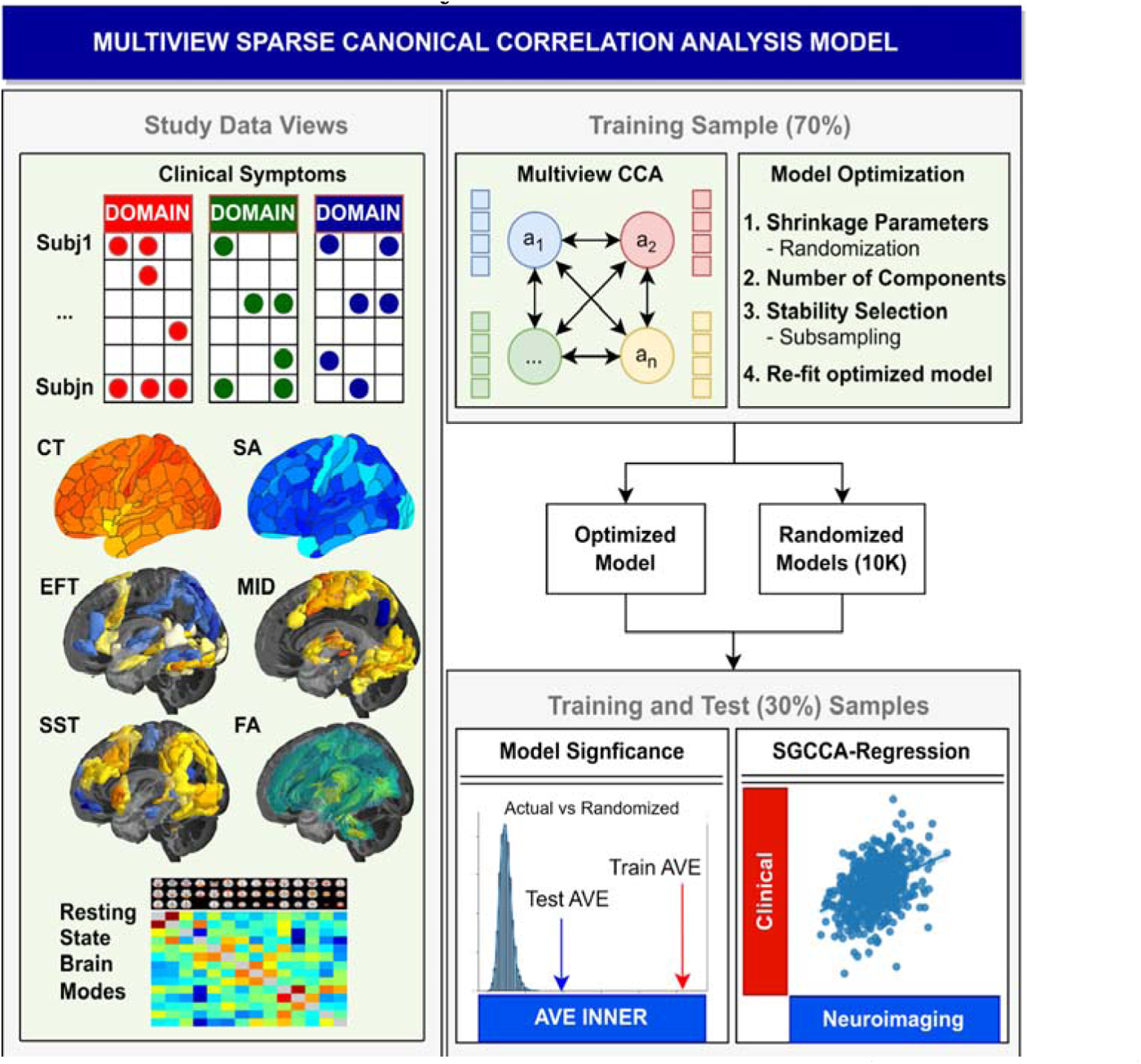
Development of sparse generalized canonical correlation analysis (SGCCA) model in the IMAGEN study. The SGCCA model incorporates eight distinct datasets (called data views), consisting of both clinical assessments and neuroimaging modalities, from the IMAGEN study. This model is built using 70% of the participants as the training dataset, while the remaining 30% form the test group. Canonical correlation analysis is the method employed, which utilizes cross-covariance matrices of two or more sets of data views to identify linear combinations (or components) that have maximal correlation. The training data serves several crucial purposes: firstly, for optimizing the model’s parameters, including shrinkage parameters (sparsity); secondly, for determining the suitable number of components; and lastly, for performing stability selection (for details refer to additional methodology). After establishing the optimal model parameters, the training data are refitted accordingly. Furthermore, ten thousand randomized models are generated by permuting subjects among each training data view. This allows us to evaluate the significance of the model within both the training and test datasets for each component. In the training data, the inner average variance explained (AVE) of the actual model is ranked and compared to the inner AVE of the randomized models. Similarly, the test data are fitted to both the actual and randomized models, and their inner AVEs are compared. Last, regression of the data view components is conducted, with clinical component scores as dependent variables and neuroimaging scores as independent variables. This entire process is repeated in both the training and test samples for each of the significant components.

**Figure 2.**
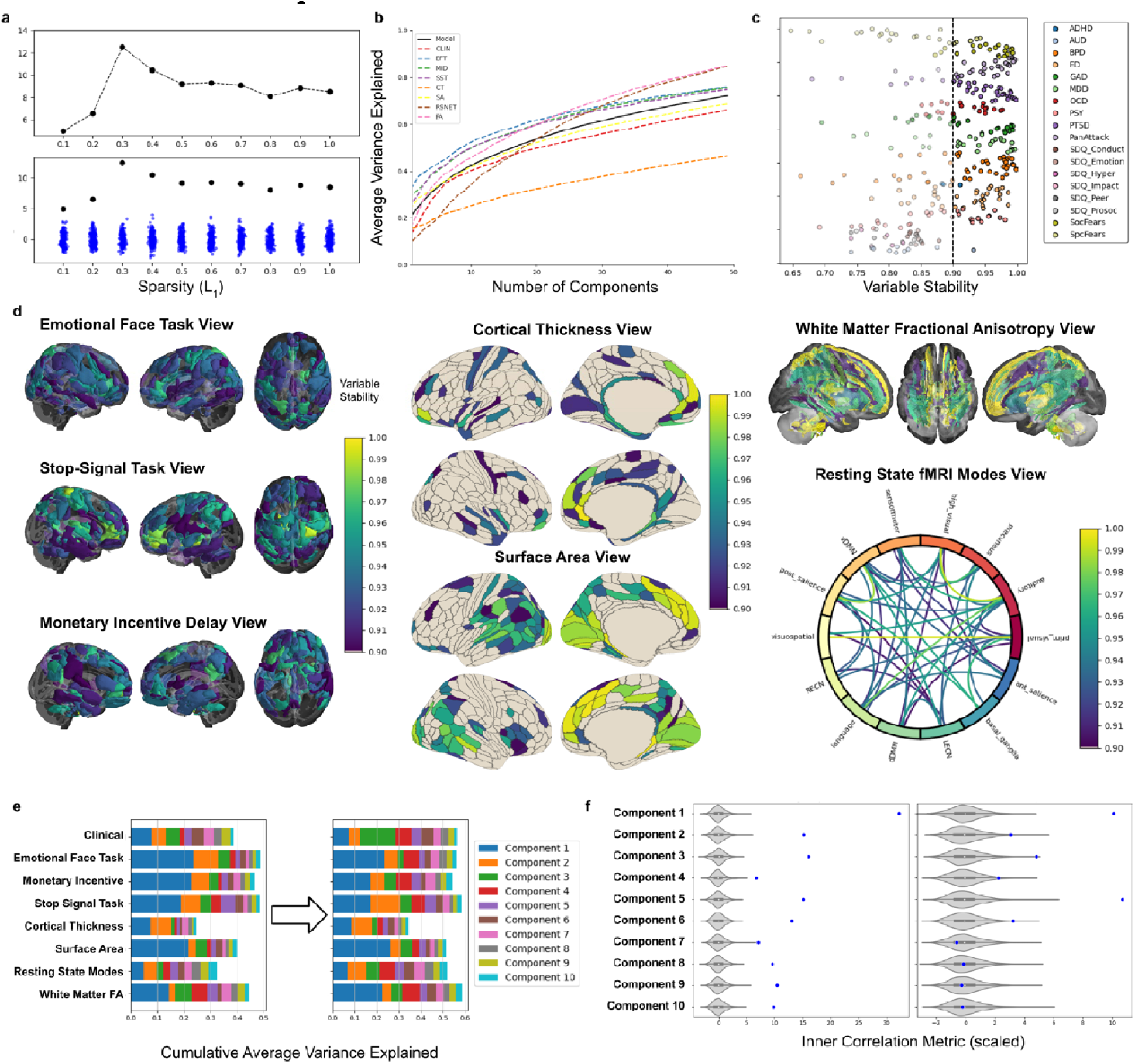
Sparse generalized canonical correlation analysis (SGCCA) model optimization, variable selection, and optimized model assessment. **a,** Line plots depict the z-scores (top) and the scaled factorial objective function (bottom), which measures covariance among the data views (black dots) and permuted data views (blue dots) for different values of L_1_ sparsity (λ_1_) ranging from 0.1 to 1.0 in intervals of 0.1. The highest z-score (z = 12.6) was achieved at λ_1_ = 0.3, indicating the optimal sparsity. **b**, Cumulative average variance explained (AVE) plotted against the number of components (x-axis) for the SGCCA model with 50 components. The solid black line represents the total model, and the dotted colored lines represent each data view. The selection of ten informative components (approximate elbow of the model curve) explains 40.4% of the cumulative variance among all data views. **c,** Stability selection by 10000 subsamples (random sampling 50% of the training data without replacement) retained the clinical items. A vertical dotted line separates variables kept in the clinical data view. **d,** Stability selection by 10000 subsamples (random sampling 50% of the training data without replacement) retained the neuroimaging data-views that appeared in 90% of the subsampled SGCCA models. Colored regions indicate brain regions used in the optimized generalized canonical correlation analysis that appeared in 90% of the subsampled SGCCA models. **e,** Barplots of the AVE for each data view aggregated along the x-axis for each component represented by different colors in the initial model (all variables, components = 10, and λ_1_ = 0.3) and the final optimized model (stability selected variables, components = 10, and λ_1_ = 1.0) in the training data. **f,** Violin plots showing the optimized permuted models for the inner average variance explained (AVE) in the training data and test sample. The AVE of the actual model are blue dots. Using the selected variables, our actual model and 10000 permuted (null) models were created. The components are considered significant if the actual model AVE is greater than 95% of the permuted models. All components in the training model were significant, whereas only the first six components were significant in the training data (p_permuted_ < 0.05).

### Establishing six independent psychopathology components

In the training data, all ten canonical components we investigated were significant (Z = 4.4 to 31.9, AVE_inner_ = 0.025 to 0.037, p_permuted_ < 1.0 x 10^-4^, Figure 2f). In the test data, the first six models remained significant (Z = 1.8 to 10.3, AVE_inner_ = 0.008 to 0.017, p_permuted_ = 0.048 to p_permuted_ < 1.0 x 10^-4^, Figure 2e). Since the first six components were significant for the model’s inner canonical correlation, we consider these to be components of interest. For an overview of the contribution of composition of individual clinical items to psychopathology scores for the six components of interest, we calculated the mean DAWBA clinical subdomains, the AUDIT, and SDQ subscales for the structural coefficients (correlation between each psychopathology score and clinical items). Based on these values, we categorized the psychopathology scores as: mania symptoms, depressive symptoms, anxiety symptoms, stress symptoms, eating pathology symptoms, and fear symptoms of components one to six, respectively (Figure 3).

**Figure 3.**
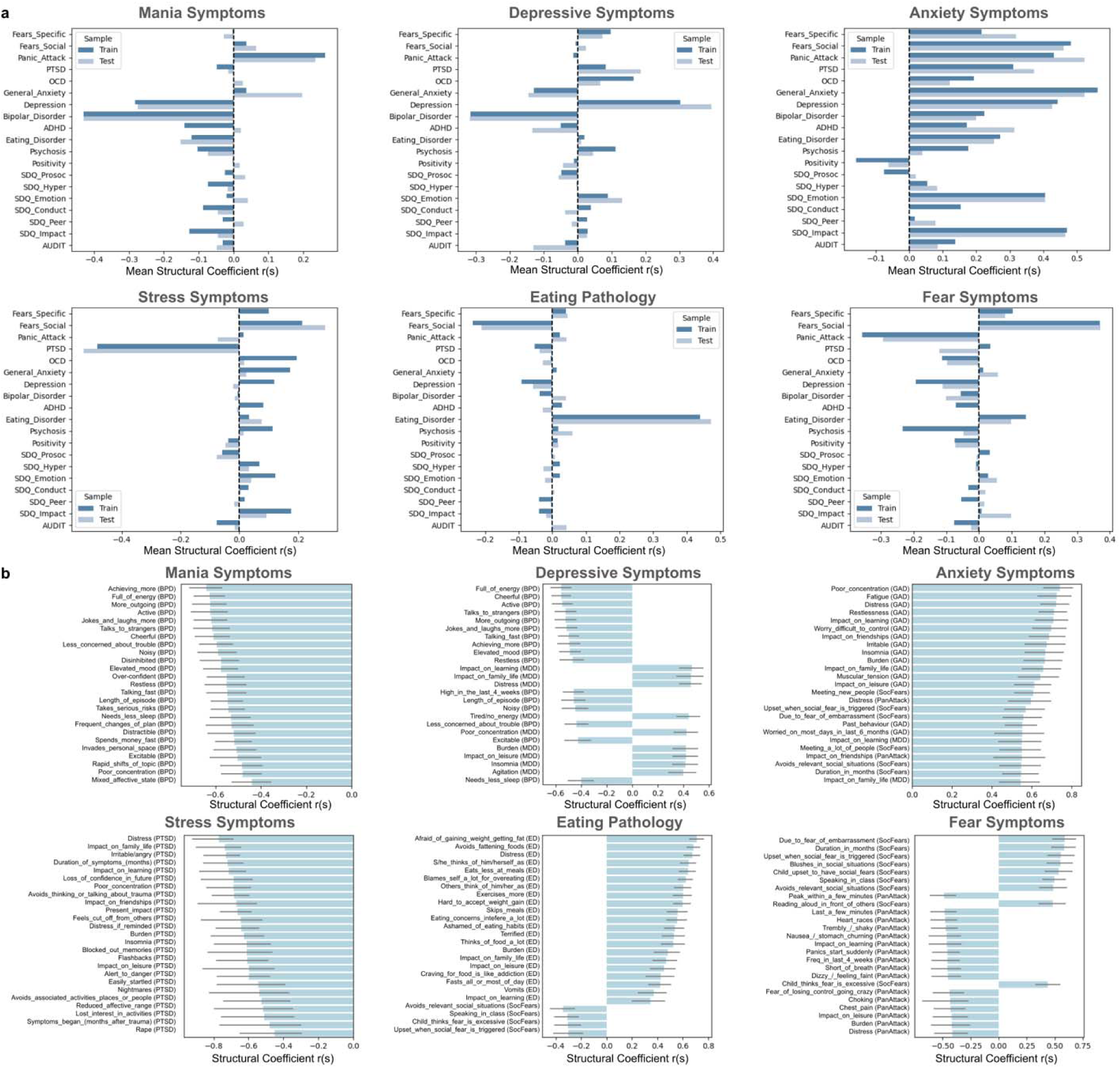
The clinical contribution to each psychopathology component. **a,** The mean structural coefficients of the significant model components among DAWBA clinical sections, the AUDIT, and SDQ subscales. The structural coefficients were calculated based on the correlation between the clinical component scores and the full set of clinical variables. The mean structural coefficients for the training and test sample are on the x-axis and can range from -1.0 to 1.0. Each item represents the mean of each DAWBA clinical section’s structural coefficients, the mean of SDQ structural coefficients, and the mean of the AUDIT structural coefficients. **b,** The top 25 items loadings are plotted according to absolute value of structural coefficients. The question from the clinical battery is on the y-axis with its corresponding section in parentheses, and the loading is on the x-axis. All items are significant (p_FDR_<0.05) after 10,000 bootstraps. ADHD, attention deficit hyperactivity disorder, AUDIT, alcohol use disorders identification test, BPD, bipolar disorder; ED, eating disorder; GAD, general anxiety disorder; MDD, major depressive disorder; OCD, obsessive compulsive disorder; PanAttack, panic attack; PTSD, post-traumatic stress disorder; SDQ, strength and difficulties questionnaire; SocFear, social fears.

### Neuroimaging contribution of the psychopathology scores

The primary study objective was to redefine psychiatric nosology based on the joint relationship between psychopathology and multimodal brain neuroimaging. The first crucial step is to evaluate which of the neuroimaging scores were contributing to psychopathology scores for each component of interest. Using SGCCA-regression, we found that each of the six symptom component scores predicted their corresponding neuroimaging components scores in both the training and test samples after Bonferroni correction for six multiple comparisons (Figure 1, p<0.0083). These associations are unsurprising in the training sample because SGCCA model optimizes variance among the clinical and neuroimaging variates; whereas, replicating the same association in the test data validates the prediction of clinical variates by the neuroimaging variates since the test data constitutes only independent transformed scores from the SGCCA model. These results were further validated in the independent cross-disorder STRATIFY/ESTRA sample composed of healthy controls and patients with major depressive disorder, alcohol use disorder, anorexia nervosa, and bulimia nervosa (Figure 4).

**Figure 4.**
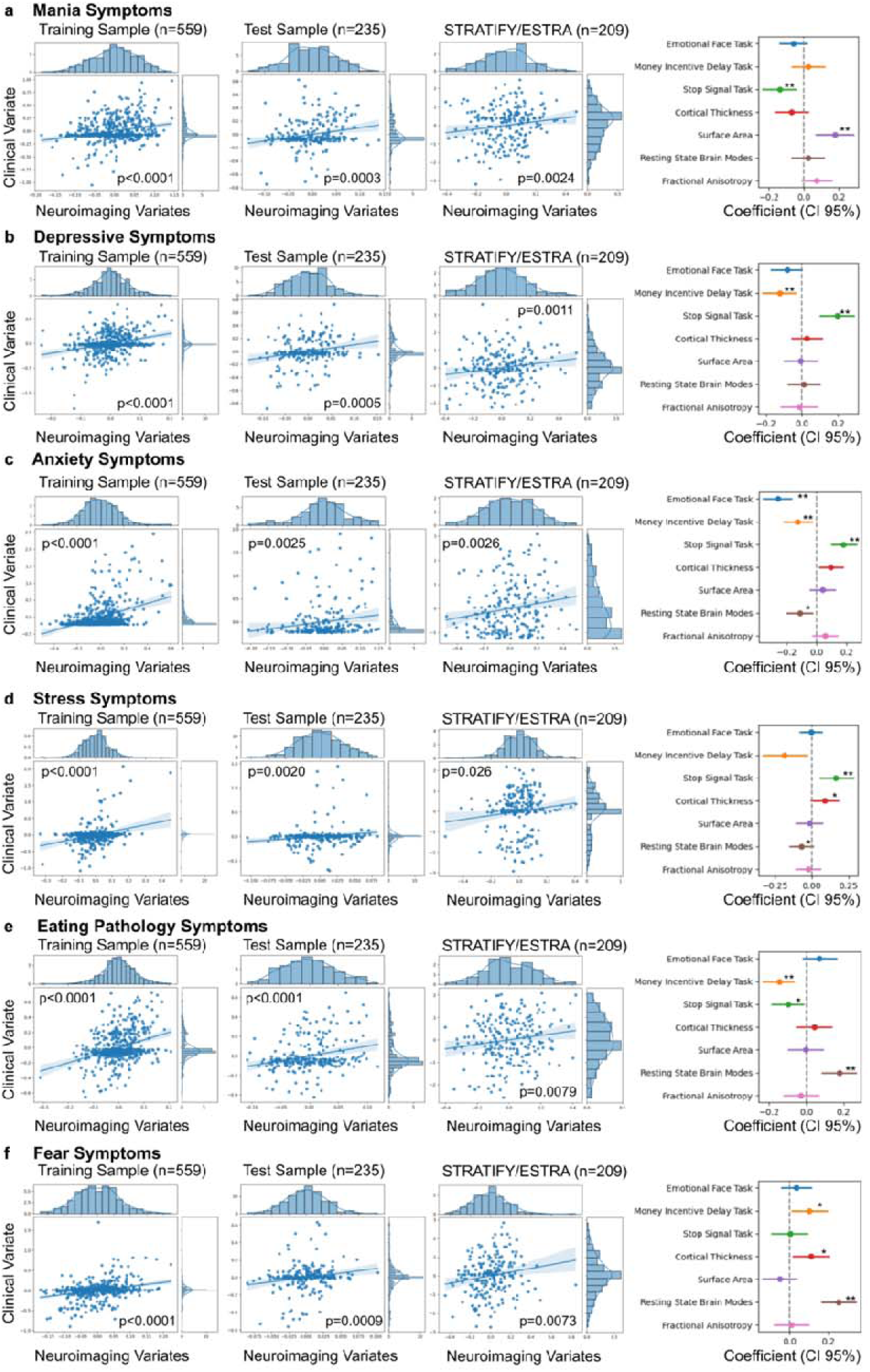
Neuroimaging features contribution to the psychopathology symptoms for each significant component. **a-f,** SGCCA-regression of the variates for the training, test IMAGEN samples and the STRATIFY/ESTRA sample with the psychopathology scores as the response variable and the neuroimaging predictor scores for: the emotional face task, the monetary incentive delay task, the stop-signal task, cortical thickness, surface area, the resting-state brain modes connectivity, and white matter fractional anisotropy. Left, regression plots and histograms of the variate psychopathology scores as the response variable and the neuroimaging predictor scores for: the emotional face task, the monetary incentive delay task, the stop-signal task, cortical thickness, surface area, the resting-state brain modes connectivity, and white matter fractional anisotropy. Right panel, bar charts of the model coefficients. Each regression model underwent 10000 bootstraps to determine the confidence interval and significance. The horizontal line represents the 95% confidence interval (2.5% to 97.5%). The psychopathology scores from components of interest were: (**a)** mania symptoms (Training Sample: r = 0.26 [0.18-0.33], Test Sample: r = 0.22 [0.10-0.35], STRATIFY/ESTRA: r = 0.19 [0.07-0.31]), (**b)** depressive symptoms (Training Sample: r = 0.30 [0.20-0.38], Test Sample: r = 0.22 [0.09-0.35], STRATIFY/ESTRA: r = 0.19 [0.04-0.33]), (**c)** anxiety symptoms (Training Sample: r = 0.40 [0.31-0.48], Test Sample: r = 0.17 [0.14-0.36], STRATIFY/ESTRA: r = 0.19 [0.06-0.30]), (**d)** stress symptoms (Training Sample: r = 0.32 [0.19-0.43], Test Sample: r = 0.14 [0.05-0.23], STRATIFY/ESTRA: r = 0.12 [0.01-0.22]), (**e)** eating pathology (Training Sample: r = 0.34 [0.25-0.42], Test Sample: r = 0.26 [0.15-0.37], STRATIFY/ESTRA: r = 0.15 [0.12-0.34]), and (**f)** fear symptoms (Training Sample: r = 0.31 [0.25-0.42], Test Sample: r = 0.18 [0.15-0.35], TRATIFY/ESTRA: r = 0.12 [0.12-0.33]). * p_bootstrap_ <0.05 ** p_bootstrap_ <0.0083.

Among the model coefficients, only cortical surface area and stop-signal stop success fMRI contrast coefficients were significant suggesting that these are primary MRI phenotypes predicting mania symptoms (Figure 4). The depressive psychopathology score was associated with monetary incentive delay large-win versus no-win fMRI contrast and stop-signal task coefficients. The anxiety symptoms score was associated with all the respective fMRI scores (Figure 4). The emotional face task angry-control contrast had the highest contribution to this association along with the monetary incentive delay task, the stop signal task, and resting-state brain modes (Figure 4). Stress symptom scores significantly associated with the stop signal task (Figure 4). Eating pathology scores had the strongest association with fMRI brain modes in addition to the monetary incentive delay task. Last, the fear symptom score was only significantly associated with resting state brain modes (Figure 4).

We tested the stability of the model by assessing consistency of the SGCCA-regression models for the six components at baseline assessment at 14 years and follow-up at 19 years by applying the SGCCA model at these time-points. In general, the previous associations observed at age 22 were also consistent at ages 14 and 19 (Supplementary Figure 3; p_bootstrap_ < 0.05), except for the manic component at age 14 in the training and test samples (p_bootstrap_ > 0.05) and the panic attack component at age 19 in the training sample (p_bootstrap_ > 0.05).

### Linked features of psychopathology and multimodal MRI modalities

Next, we asked which variables are driving the association between psychopathology scores and neuroimaging modality scores for each component (Figure 4). This step is important for identifying which clinical items are most linked to brain regions - information that could be used to develop a parsimonious model to be applied in a clinical setting.

The psychopathological variables contributing to the mania symptoms were primarily negative loading from DAWBA questions from the bipolar disorder section (P_FDR_ < 0.05; Figure 3b). The stop-signal task negatively loaded strongly in areas involved in frontoparietal executive function which mirrored the surface area loading in the dorsolateral prefrontal cortex, anterior cingulate and inferior parietal cortex (P_FDR_ < 0.05; Figure 5a). These regions play an important role in cognitive control, including attention and response inhibition and are altered in bipolar patients and their relatives ^12,13^.

The clinical loadings for our depressive symptom score were observed in opposing directions for bipolar and major depressive disorder symptoms. The score was negatively loaded with bipolar items related to “full of energy”, “more active”, “elevated mood”, and positively loaded with depressive items such as “miserable daily”, “impact of depression”, “tired or low energy”, “feelings of worthless guilt” (P_FDR_ < 0.05; Figure 3b). Since the mania symptoms score is orthogonal to this score, we consider depressive to be the primary psychopathological loadings. The monetary incentive delay and stop-signal tasks both loaded in anterior and posterior cingulate cortex, in opposing directions (P_FDR_ < 0.05; Figure 5b). The anterior cingulate modulates emotion-driven behaviors and higher cognitive function ^14^; whereas, the posterior cingulate cortex is integral to the default mode network and involved in memory retrieval and planning ^15^ suggesting involvement of cognitive and emotional processing .

The anxiety symptom score positively loaded most on questions related to poor concentration, impact on learning, and distress from the general anxiety disorder DAWBA section with additional loadings from the social fears, depression, and panic attack items (P_FDR_ < 0.05; Figure 3b). The insula and superior temporal gyrus were common loadings for the emotional face, monetary incentive delay, and stop-signal tasks with the latter also loading in the medial prefrontal cortex. The task loadings would suggest a common involvement of salience and ventral attention networks (P_FDR_ < 0.05; Figure 5c). Similarly, in the resting-state brain modes loadings there was a strong negative correlation among the anterior salience and visual networks (P_FDR_ < 0.05; Figure 5c).

The stress symptom score was primarily negatively loaded to post-trauma stress disorder DAWBA questions (P_FDR_ < 0.05; Figure 3b). The stop-signal task score had significant loading in the dorsal anterior cingulate cortex, insula, as well as the pre- and post-central gyrus suggesting an involvement with default mode salience networks (P_FDR_ < 0.05; Figure 5d). Significant cortical thickness loading was in the anterior and posterior cingulate, orbitofrontal, dorsolateral prefrontal, and insular cortices (P_FDR_ < 0.05; Figure 5d).

The eating pathology score loaded more on DAWBA questions related to bulimia nervosa than anorexia nervosa; although, both were present (P_FDR_ < 0.05; Figure 3b). Loadings in the striatum, medial prefrontal cortex, and medial temporal cortex were present for the monetary incentive delay task suggesting an involvement of limbic and anterior salience networks (P_FDR_ < 0.05; Figure 5e). There was a negative correlation loading between the high visual and language networks (medial temporal), and the language network positively correlated with the ventral default mode network (P_FDR_ < 0.05; Figure 5e).

The clinical loadings for the fear symptom score were split between social fears and panic attack items, but since this component is orthogonal to the previous anxiety and social fear component, we consider panic attack symptoms to be the main clinical loading (P_FDR_ < 0.05; Figure 3b). The dorsal default mode network negatively correlated with both the visual and sensorimotor networks in the resting state brain modes (P_FDR_ < 0.05; Figure 5f).

**Figure 5.**
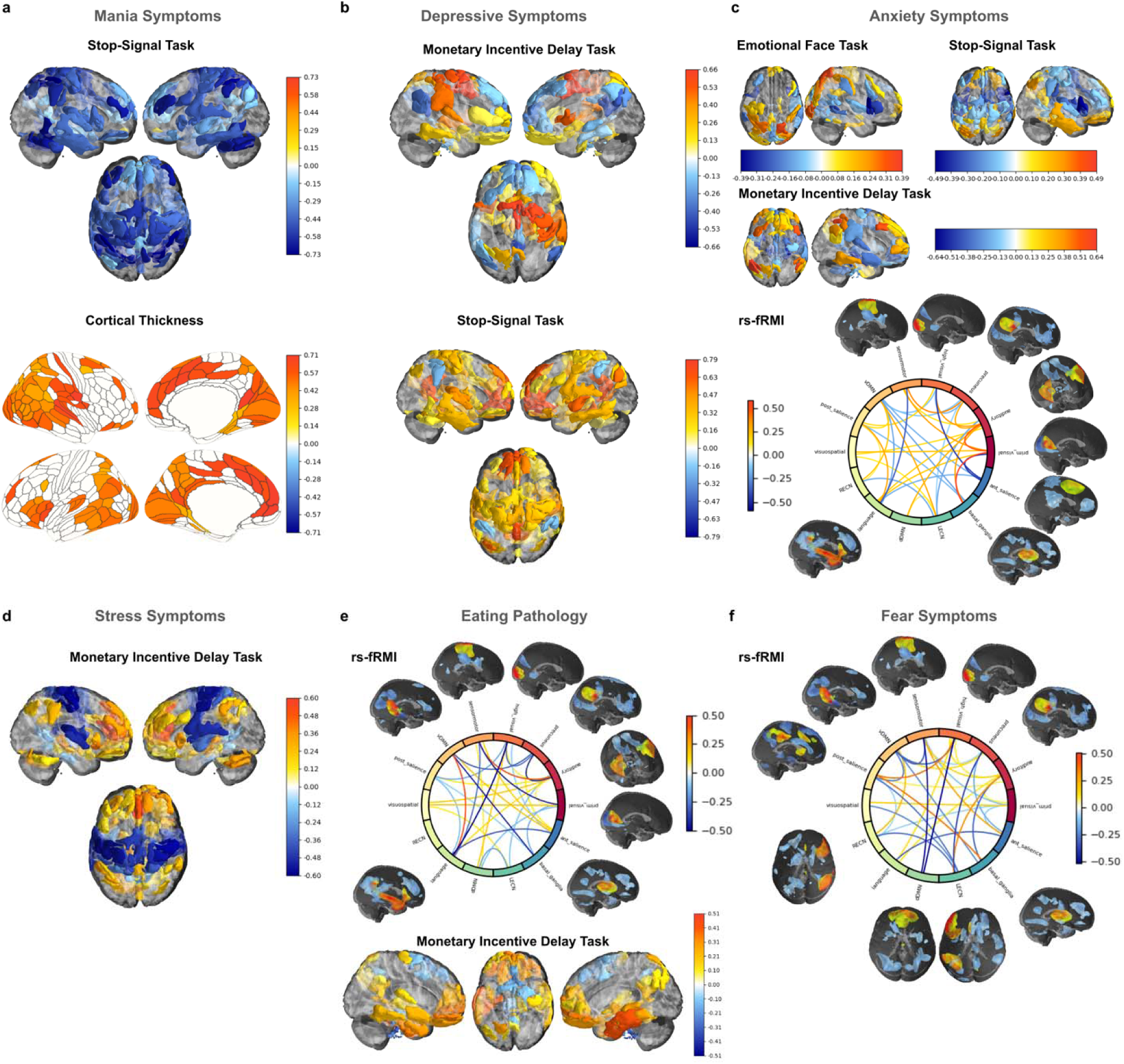
Neuroimaging loadings for each psychopathology score. **a-f,** Significant loadings (structural coefficients; r_s_) for each score are shown using 10000 bootstraps and after accounting for false discovery rate (P_FDR_ < 0.05). Colors ranging from red to dark blue denote significant positive and negative r_s_ values, respectively. The psychopathology components of interest were **(a)** mania symptoms: stop-signal task and cortical thickness scores, (**b)** depressive symptoms: monetary incentive delay task and stop-signal task scores, **(c)** anxiety symptoms: emotional face task, stop-signal task, monetary incentive delay task, and resting state brain modes (rs-fMRI) scores **(d)** stress symptoms: monetary incentive delay score, **(e)** eating pathology: monetary incentive delay and rs-fMRI scores, and (**f)** fear symptoms: rs-fMRI scores.

## Discussion

We have developed a framework for psychiatric nosology that is based upon quantitative neurobiological measures. By constructing symptom groups according to shared structural and functional neuroimaging features across seven modalities, we have provided mechanistic characterization and identified targets for therapeutic intervention. As our clinical characterization is based upon existing ICD-10/DSM-5 symptoms that were simply reassembled according to a shared underlying biology, we have preserved the clinical experience accumulated in existing psychopathological characterizations while optimizing them for neurobiological prediction. This approach facilitates application of our framework in clinical research studies and psychiatry services. Its clinical relevance is further augmented by the surprising fact that the data-driven canonical correlates that we identified closely mimic existing psychiatric diagnoses, thus requiring merely an adaptation of individual symptoms to obtain a refined definition of mental disorders that is characterized by underlying biological mechanisms.

The ability to link major psychiatric symptoms, including mania, depression, anxiety, stress, eating pathology, and fear, to distinct neuroimaging modalities not only helps with biological understanding of those symptoms but also provide important insights to the manifestation of mental disease since each components has a quantifiable, biological measure that is independent from the other components. The model demonstrates strong predictive stability as it replicates in both the independent test dataset and the cross-disorder STRATIFY/ESTRA sample. While the model was developed among the IMAGEN participants, who are young adults at the age of 23, the clinical associations largely remained consistent at ages 14 and 19 suggesting that the neuroimaging variables may serve as early markers of severe symptoms. The regression analysis of the six clinical components reveals distinct associations with neuroimaging modalities; although, regions involved in executive functioning and the default mode network consistently emerge.

Previous neuroimaging-CCA studies have identified between one and three significant components ^9,16–19^. The fact that our study describes six components enhances discrimination and renders our framework useful for diagnostic classification. In our framework, diagnostic specificity and precision are enhanced by the direct correlation of symptom groups with brain structure and function. Diagnosis relies not only on symptom counts but also on the precise weighting of individual symptoms within each symptom group. In this way, scores for each symptom can be derived providing a quantitative estimate of comorbidity to guide treatment decisions. Furthermore, the framework provides a rational way for escalating diagnostic examinations. Whereas symptom scores might suffice for treatment decisions in uncomplicated cases, targeted neuroimaging analyses will provide more precise quantitative assessments of disease mechanisms and objective measures of disease course in clinic and research.

The data-driven neuroimaging characterization of psychopathology provides new insight into multimodal brain relationships and their symptom-specific liability regions. The mania score (component 1) was associated with a novel structure-function relationship, involving activation during the stop-signal task and joint surface area in overlapping regions including the medial frontal gyrus, inferior frontal gyrus, insula, inferior parietal cortex, caudate, and putamen. By demonstrating the contingency of functional activation of these brain areas implicated in behavioral inhibition ^20^ on the regional surface area, our finding provides a more refined understanding of the mechanisms that could contribute to impaired inhibitory control in mania^21,22^.

For the depressive score (component 2) overlapping activations during both tasks were observed in the dorsolateral prefrontal cortex, medial prefrontal cortex, posterior cingulate cortex, precuneus, and limbic regions including the hippocampus and amygdala. The novelty of this finding lies in the identification of a fronto-limbic brain network involved in top-down control and emotion integration ^23–25^ that is specific to depressive symptoms and distinct from the mania component 1.

The anxiety score (component 3) was the only component broadly associated with multiple ICD-10/DSM-5 diagnostic categories with individual items primarily related to anxiety. It was associated with all functional neuroimaging modalities - but not the structural modalities - in the amygdala, thalamus, and insula with involvement of the anterior salience network. Anxiety symptoms are frequently present in psychiatric disorders, particularly in internalizing and thought disorders ^26–28^. The anxiety score contains clinical and multimodal brain relationships that are remarkably similar to those of a recently described transdiagnostic neuropsychological factor, in particular the involvement of the medial prefrontal cortex, insula, and thalamus ^29^. However, since we identified six transdiagnostic psychopathology components (Figure 3, Figure 4) with unique multimodal associations, we consider component 3 as a distinct diagnostic entity as opposed to a transdiagnostic vulnerability factor.

The stress score (component 4) was associated with the stop-signal task with the strongest positive loading in the anterior cingulate cortex consistent with hyperactivation in this region associated with emotional reactivity and vigilance. The clinical items contributing to the stress scores were predominately related to post-traumatic stress disorder, suggesting a link to this score and prior trauma. This relationship is bolstered by the pivotal role of the anterior cingulate cortex in emotional reactivity and post-traumatic stress disorder ^30,31^.

The eating pathology score (component 5) was strongly associated with the resting state brain modes and particularly connectivity between the ventral default mode, basal ganglia, and temporal networks. Both functional and structural associations with the temporal lobe have been reported in bulimia nervosa, where these networks are thought to be linked to social behavior and emotional stimuli ^32–34^.

Fear symptoms (component 6) was only associated with resting state brain modes, particularly with respect to connectivity in the dorsal default mode and left executive control networks suggesting neural mechanisms underlying deficits in cognitive control during experiences of fear ^35^.

While our findings are supported by the current literature of psychiatric disorders. The associations with symptoms we describe are independent of each other. As with the symptom scores themselves, they mimic neuroimaging findings in psychiatric diseases. The key difference in our neuroimaging findings is that they are specific which would make them better biomarkers for clinical neuroimaging studies.

Our study has certain limitations. The neuroimaging predictors for the clinical components were modeled using a naturalistic sample, minimizing potential confounds from psychiatric treatment, such as medications, but likely missing out on psychiatric disorders with a lower prevalence (such as schizophrenia) or a different age distribution (such as dementia). To capture these conditions, samples enriched for the target disorder are required. Similarly, other neuroimaging tasks interrogating different cognitions and other modalities known to influence mental illness, such as -omics and environmental factors need to be considered in future studies. For all these refinements, an analytical strategy similar to the one presented can be employed. Hence, our approach provides a framework for the development of a psychiatric nosology that is both flexible and scalable.

In conclusion, jointly linking psychiatric symptoms to multimodal brain features lays the groundwork for a brain-informed psychiatric nosology, with immediate implications to treatment decision, and novel therapeutic development. By identifying symptom groups that are directly linked to quantifiable neurobiological measures, our approach enables the development of interventions that precisely target causal mechanisms of disorder. Weighting of each individual symptom enables quantitative assessment of comorbidity, further increasing the precision of interventions. Together, our results suggest the feasibility of data-driven nosologic frameworks and demonstrate their potential to bridge the gap between psychiatric neuroscience and clinical treatment of mental disorders.

## Online Methods

### Participants

The analysis was carried out on 794 (366 male and 428 female) adolescents from the population-based, longitudinal child development cohort, IMAGEN, with neuroimaging assessments 14, 19, and 23 years with an additional psychological assessment at 16 years ^36^. Additionally, 212 (29 male and 150 female) participants from the cross-disorder STRATIFY/ESTRA clinical cohorts were included, whose assessments were collected in tandem with the final follow-up of the IMAGEN at age 23. Details of the cohorts are available in additional methodology, Supplementary Tables 1-3. We analyzed participants with complete clinical assessments of The Development and Well-Being Assessment (DAWBA), Strengths and Difficulties Questionnaire (SDQ), Alcohol Use Disorders Identification Test (AUDIT), as well as quality-controlled neuroimaging data including T1-weighted structural MRI, diffusion weighted images, resting state and task-based functional MRI (fMRI).

### Clinical characterization

Clinical psychiatric symptoms were assessed using the single items from the DAWBA^37^, AUDIT ^38^, and SDQ ^39^. DAWBA screening questions have previously been used to define subthreshold clinical symptoms in neuroimaging studies of psychopathology ^37,40^. The SDQ was also used in the present investigation, as this questionnaire contributes to the assignment of diagnostic status in the DAWBA ^39^. The AUDIT questionnaire was used to screen for potential harmful drinking and identify mild dependence ^38^. For all questionnaires, we were interested in the symptoms rather than defined diagnostic criteria; therefore, we considered all items in these questionnaires rather than only the entry items. We posit that this strategy allows us to describe precise interrelationships among different categories of questions. However, a consequence using all items in structural interviews means that we treat questions that have not been asked as having no symptoms. For example, if a subject answered no to the entry question, “Overall, do you particularly fear or avoid social situations that involve a lot of people, meeting new people, or doing things in front of other people?”, we assumed that they also do not feel afraid of meeting new people in the subsequent follow-up question. In total, there were 335 items among all questionnaires after two items from the DAWBA were dropped because they had zero variance across the training sample.

### Neuroimaging procedures

Magnetic resonance imaging acquisition and processing were performed according to IMAGEN guidelines, and the details are available in the additional methodology.

### Sparse Generalized Canonical Correlation Analysis (SGCCA)

Details of the SGCCA model and its derivation have been described elsewhere ^41,42^, and sparse canonical correlation analysis is common among neuroimaging analyzes ^16,41,42^. Details of our model parameters, optimization are available in the additional methodology. SGCCA is a method that uses cross-covariance matrices of two or more sets of vectors (or data views) to find the linear combinations (or components) of these data views (clinical or neuroimaging data) that have maximum correlation with each other using gradient descent. We include all data views into a unified model; thus, describing multimodal functional, structural, and diffusion MRI relationships in the context of cross-disorder symptom scores (Figure 1).

### Statistical analysis

After model optimization, we employed another permutation scheme to compare the canonical correlates of the real data SGCCA model and rank them against 10,000 permutations of permuted (null) SGCCA models (Figure 1). We calculated the average inner variance explained (AVE_inner_), represented by the mean of canonical correlations. The real data canonical variates were then ranked against the null models to determine their permuted p-values. To rule out possible overfitting of the model, we performed an additional assessment of significance using independent test data. We utilized the SGCCA model derived from the real training data and applied it to the test data. This involved taking the dot product of the training model coefficients and the test data views to calculate their canonical correlates. Subsequently, we compared the predicted canonical correlates in the test data to the predicted canonical correlates obtained from the 10000 permuted SGCCA models, rank ordering them for significance.

We considered a component to be of interest if the AVE_inner_ was significant in both the training data and the test data. We did not assess the significance of the model components alone, as the average inner canonical correlations would encompass correlations among all data view scores equally. Our primary focus was on the association between the clinical components and the other seven neuroimaging data views. We employed a combined regression and canonical correlation analysis SGCCA approach, which is a widely used and well-established method in multiview canonical correlation analyzes ^16,42,43^. By utilizing regression alongside SGCCA, the combined approaches allowed us to specifically examine the association between the clinical components and the neuroimaging data views of interest. We conducted SGCCA-regression analyses to investigate the relationships between the latent clinical component (response variable) and the latent neuroimaging components (predictor variables), separately in the training and test IMAGEN data. The identical method was applied at the 14 and 19 timepoints of the IMAGEN training and test subjects as well as STRATIFY/ESTRA. To determine the significance of the model and the coefficients, we employed 10,000 bootstraps. Components of interest were deemed significant only if the regression model showed significance in both the training and test data.

Last, we assessed the model loadings in the training data, which represent the correlation between the view components and their respective variables. These loadings indicate the amount of variance explained by each variable. The significance and confidence intervals of the loadings were evaluated using 10,000 bootstraps and Benjamin-Hochberg false discovery rate correction (FDR)^44^.

## Reporting summary

Further information on research design is available in the Nature Portfolio Reporting Summary linked to this article.

## Data availability

IMAGEN data are available from a dedicated database at https://imagen-project.org. STRATIFY/ESTRA data are available from the IMAGEN database at https://stratify-project.org.

## Code availability

The code that supports the findings of this study is available on GitHub at https://github.com/trislett/sgcca-psychiatry-nosology.git.

## Acknowledgments

This work received support from the following sources: the European Union-funded Horizon Europe project ‘environMENTAL’ (101057429) and co-funding by UK Research and Innovation under the UK Government’s Horizon Europe funding guarantee (10041392 and 10038599); the Horizon 2020-funded European Research Council Advanced Grant ‘STRATIFY’ (695313); the German Research Foundation (COPE; 675346); the European Union-funded FP6 Integrated Project IMAGEN (Reinforcement-related behaviour in normal brain function and psychopathology) (LSHM-CT-2007-037286), theLMedicalLResearch Council and Medical Research Foundation (grants MR/R00465X/1 and MRF-058-0004-RG-DESRI: ‘ESTRA: Neurobiological underpinning of eating disorders: integrative biopsychosocial longitudinal analyses in adolescents’; MR/S020306/1 and MRF-058-0009-RG-DESR-C0759: ‘Establishing causal relationships between biopsychosocial predictors and correlates of eating disorders and their mediation by neural pathways’) the Medical Research Council (grant MR/W002418/1: ‘Eating Disorders: Delineating illness and recovery trajectories to inform personalized prevention and early intervention in young people (EDIFY)’ and the National Institute for Health and Research (NIHR) Biomedical Research Centre (BRC) and Maudsley NHS Foundation Trust (SLaM). The Human Brain Project (HBP SGA 2, 785907, and HBP SGA 3, 945539), the National Key R&D Program of Ministry of Science and Technology of China (MOST 2023YFE0199700). The Medical Research Council Grant ’c-VEDA’ (Consortium on Vulnerability to Externalizing Disorders and Addictions) (MR/N000390/1), the National Institute of Health (NIH) (R01DA049238, A decentralized macro and micro gene-by-environment interaction analysis of substance use behavior and its brain biomarkers), the Bundesministerium für Bildung und Forschung (BMBF grants 01GS08152; 01EV0711; Forschungsnetz AERIAL 01EE1406A, 01EE1406B; Forschungsnetz IMAC-Mind 01GL1745B), the Medical Research Foundation (MRF-058-0014-F-ZHAN-C0866) and the National Institutes of Health (NIH) funded ENIGMA (grants 5U54EB020403-05 and 1R56AG058854-01), NSFC grant (82150710554).

Further support was provided by grants from: - the ANR (ANR-12-SAMA-0004, AAPG2019 - GeBra), the Eranet Neuron (AF12-NEUR0008-01 - WM2NA; and ANR-18-NEUR00002-01 - ADORe), the Fondation de France (00081242), the Fondation pour la Recherche Médicale (DPA20140629802), the Mission Interministérielle de Lutte-contre-les-Drogues-et-les-Conduites-Addictives (MILDECA), the Assistance-Publique-Hôpitaux-de-Paris and INSERM (interface grant), Paris Sud University IDEX 2012, the Fondation de l’Avenir (grant AP-RM-17-013), the Fédération pour la Recherche sur le Cerveau; the National Institutes of Health, Science Foundation Ireland (16/ERCD/3797), U.S.A. (Axon, Testosterone and Mental Health during Adolescence; RO1 MH085772-01A1) and by NIH Consortium grant U54 EB020403, supported by a cross-NIH alliance that funds Big Data to Knowledge Centres of Excellence. This paper represents independent research partly funded by the NIHR Maudsley Biomedical Research Centre at South London and Maudsley NHS Foundation Trust and King’s College London. The views expressed are those of the authors and not necessarily those of the NIHR or the Department of Health and Social Care or the European Union, the European Health and Digital Executive Agency (HADEA) or UKRI.

We also thank the additional raters for the DAWBA-SDQ: Patricia Conrod, Maren Struve, Jani Penttilä, Viola Kappel, Yvonne Grimmer, Tahmine Fadai, Corinna Insensee, Andreas Becker, Andreas Becker-Isensee.

## Ethics declarations

Dr. Banaschewski served in an advisory or consultancy role for Lundbeck, Medice, Neurim Pharmaceuticals, Oberberg GmbH, Shire. He received conference support or speaker’s fee from Lilly, Medice, Novartis and Shire. He has been involved in clinical trials conducted by Shire & Viforpharma. He received royalties from Hogrefe, Kohlhammer, CIP Medien, Oxford University Press. The present work is unrelated to the above grants and relationships. Dr. Poustka served in an advisory or consultancy role for Roche and Viforpharm and received a speaker’s fee from Shire. She received royalties from Hogrefe, Kohlhammer and Schattauer. The present work is unrelated to the above grants and relationships. The other authors report no biomedical financial interests or potential conflicts of interest.

## Contributions

T.A.L. and G.S. wrote the manuscript, and feedback and editing were provided by all authors. T.J., A.M., E.P., contributed to statistical analysis. N.V., T.B., A.L.W.B., H.F., A.G., H.G., P.G., A.H., R.B., J.-L.M, M.-L.P.M., E.A. F.N., D.P.O., H.V., T.P., L.P., M.B., M.J.B., B.M.N., A.S., Z.Z., L.R., J.W., Y.Z., S. K., C. B., M.N.S, R.W, U.S., J.S., H.W. S.D., acquired the data. A.L.W.B., L.W., H.L., R.L., E.A., contributed to neuroimaging data processing and analysis. T.A.L., G.S., A.H., J.F., T.W.R., S.D., A.M. conceptualized the study.

## Corresponding authors

Correspondence to Tristram Lett or Gunter Schumann.

