## Extended Data 1 for "A framework for a brain-derived nosology of psychiatric disorders"

*Supplementary Information*

### Additional Methodology

*Cohort assessments*

The cohorts include predominantly White, Europeans (self-described and validated using principal component analysis of population stratification informative genetic variants ^1^ from eight clinical research hospitals in: England, France, Germany, and Ireland. IMAGEN includes an extensive clinical, biological, and assessments that has been previously described ^2^. Each research site received approval from the relevant local research ethics committee and in accordance with the Declaration of Helsinki. Written consent was obtained from each participant and a parent or guardian. The Development and Wellbeing Assessment (DAWBA)^3^, Strengths and Difficulties Questionnaire (SDQ)^4^, and Alcohol Use Disorders Identification Test (AUDIT) ^5^, as well as  neuroimaging assessments performed in STRATIFY and ESTRA were nearly identical to IMAGEN’s assessment. In addition to healthy controls (n=26), STRATIFY/ESTRA also contains participants meeting diagnostic criteria for patients diagnosed with: attention deficit hyperactivity disorder (ADHD; n=1), alcohol use disorder (AUD; n=64), major depressive disorder (MDD; n=78), and psychosis (n=3) (i.e., STRATIFY sample), and anorexia nervosa (AN; n=22), bulimia nervosa (BN; n=19) (i.e., ESTRA sample). Due to the small sample size of the ADHD and psychosis groups with complete data, they were excluded from the present study.

*Image acquisition*

MRI scans were acquired from 3-Tesla scanners from different manufacturers (Siemens, Munich, Germany; Philips, Best, The Netherlands; General Electrics, Chalfont St Giles, UK; Bruker, Ettlingen, Germany) at eight different sites (King’s College, London; Sir Peter Mansfield Imaging Centre of the University of Nottingham; Trinity College Institute of Neuroscience, Dublin; the Centre de Neuroimagerie de Recherche, Paris; Charité Universitätsmedizcine Berlin; Universitätsklinikum Hamburg-Eppendorf; Zentralinstitut für seelische Gesundheit, Mannheim; and the Universitätsklinikum Carl Gustav Carus, Dresden). Acquisition protocols for sMRI, DWI, resting state fMRI (rs-fMRI), as well as monetary incentive delay (MID), emotional faces task (EFT), and stop signal task (SST) as described in previous publications ^1,2,6,7^. In brief, high-resolution anatomical MRIs were acquired, including a three-dimensional (T1-weighted; sagittal slice plane; repetition time (TR) 2.3s; echo time (TE) 2.93ms; flip angle 9°; 256×256×160 matrix; isotropic voxel size 1.1mm) magnetization prepared gradient echo sequence (MPRAGE) based on the ADNI protocol. DWI image acquisition was identical across sites using an echo planar imaging (b=0 and 32 directions with b-value 1,300 s*mm^-2^ ; axial slice plane; echo time = 104ms; 128×128×60 matrix; voxel size 2.4×2.4×2.4mm). The fMRI standardized acquisition parameters were performed using a single-shot T2*-weighted gradient-echo echo planar imaging (GE-EPI) sequence with 3.4 x 3.4 mm in-plane resolution, 164 volumes, 40 slices with a thickness of 2.4 mm with a gap of 3.4 mm, TR of 2200 ms, TE of 30 ms, a flip angle of 75°, and a Field of View of 218 x 218 mm. Volumes were acquired in sequential ascending slice order. For rs-fMRI, participants were instructed to close eyes and to not focus on a specific thought (5-6 min duration). All imaging data were visually screened for corrupted data or acquisition artifacts. For the STRATIFY sample, the identical MRI acquisitions were performed at three of the eight sights (King’s College, London; Charité Universitätsmedizcine Berlin; and University of Southampton, Southampton)

*Processing of structural images*

Cortical reconstruction was performed on all T1-weighted images using the Freesurfer image analysis suite (http://surfer.nmr.mgh.harvard.edu/). The technical details of these procedures are described in prior publications ^8–12^. In brief, this process includes motion correction and averaging of multiple volumetric T1 weighted images, removal of non-brain tissue, automated Talairach transformation, segmentation of the subcortical white matter and deep gray matter volumetric structures, intensity normalization, tessellation of the gray matter white matter boundary, automated topology correction, and surface deformation. A number of deformable procedures were performed including surface inflation, registration to a spherical atlas which is based on individual cortical folding patterns to match cortical geometry across subjects, and creation of a variety of surface-based data including maps of curvature and sulcal depth. Both intensity and continuity information from the entire three dimensional MR volume in segmentation and deformation procedures to produce representations of cortical thickness, calculated as the closest distance from the gray/white boundary to the gray/CSF boundary at each vertex on the tessellated surface ^8^.

*Processing of Diffusion Weighted Images*

Diffusion data preprocessing was performed using tools provided by the MRtrix3 software package [http://mrtrix.org](http://mrtrix.org/) (Tournier et al., 2019), including, for some preprocessing step, scripts interfacing with the external package FSL FMRIB Software Library (FSL) [https://fsl.fMRIb.ox.ac.uk/fsl/fslwiki/](https://fsl.fmrib.ox.ac.uk/fsl/fslwiki/). Diffusion images were denoised (Veraart et al., 2016), corrected for Gibbs ringing artifacts (Kellner et al., 2016), and corrected for eddy current-induced distortions and subject movements (Andersson and Sotiropoulos, 2016) using outlier replacement (Andersson et al., 2016) and a linear second level model. Then, a B1 field inhomogeneity correction was performed using the N4 algorithm as provided in ANTs (Turtison et al, 2010). The diffusion tensor model was fitted using a weighted least-squares estimator (Veraart et al., 2016) before to extract derivative diffusion metrics (FA, MD, AD and RD). Following methodological recommendations on diffusion tensor image registration ^13^ and on tract-based spatial statistic ^14^, We used a tensor-based registration with DTI-TK <http://dti-tk.sourceforge.net/pmwiki/pmwiki.php> ^15^ to align all individuals into a common space. The multiple time-point images were processed within an unbiased longitudinal framework using tensor-based registration ^16^. A first step created a within-subject template based on the multiple time point images per subject. Then a second step created a group-wise atlas based on the within-subject templates. At the end of the registration procedure, each participant’s diffusion data were normalized to the MNI standard space using the IIT human brain tensor template ^17^. Voxel-wise statistical inference of the diffusion tensor metrics was carried out using the TBSS procedure ^18^. From all previously obtained images, the mean FA image was created and thinned to create a mean FA skeleton, which represents the centers of all tracts common to the group. This skeleton was then thresholded to FA > 0.2 to keep only the main tracts. Each participant’s aligned fractional anisotropy images were then projected onto this common skeleton to minimize any residual misalignment of tracts.

*Task-based fMRI*

To assess fMRI blood-oxygen-level-dependent (BOLD) activation in responses to reward anticipation and reward outcome ^19^, the participants performed the monetary incentive delay (MID) fMRI task. In 66 ten-second trials, participants were presented with one of three cue shapes (cue, 250 ms) denoting whether a target (white square) would subsequently appear on the left or right side of the screen, and whether 0, 2 or 10 points could be won in that trial. After a variable delay (4,000-4,500 ms) of fixation on a white crosshair, participants were instructed to respond with left/right button-press as soon as the target appeared. Feedback on whether and how many points were won during the trial was presented for 1450 ms after the response. Our outcome contrast for this task was large-win versus no-win.

For fMRI BOLD response to emotional stimuli, we used the emotional face task (EFT) ^20^. Participants watched 18-second blocks of either a face movie (depicting anger or neutrality) or a control stimulus. Each face movie showed black and white video clips (200-500ms) of male or female faces. Five blocks each of angry and neutral expressions were interleaved with nine blocks of the control stimulus. Each block contained eight trials of 6 face identities (3 female). The same identities were used for the angry and neutral blocks. The control stimuli were black and white concentric circles expanding and contracting at various speeds that roughly matched the contrast and motion characteristics of the face clips. As in previous IMAGEN studies, our primary contrast was viewing angry faces (vs control) ^7^.

Last, we assess fMRI BOLD activation during response inhibition using the stop-signal task ^21^. The task was composed of Go trials and Stop trials that activate top-down attentional control particularly in the inferior frontal cortex. During Go trials (83%; 480 trials) participants were presented with arrows pointing either to the left or to the right, and subjects were instructed to make a button response with their left or right index finger corresponding to the direction of the arrow. In the unpredictable Stop trials (17%; 80 trials), the arrows pointing left or right were followed (on average 300 ms later) by arrows pointing upward and subjects were instructed to inhibit their motor responses during these trials. A tracking algorithm changes the time interval between Go signal and Stop signal onsets according to each subject’s performance on previous trials (average percentage of inhibition over previous Stop trials, recalculated after each Stop trial), resulting in 50% successful and 50% unsuccessful inhibition trials. The inter-trial interval was 1800 ms. The tracking algorithm of the task ensured that subjects were successful on 50% of Stop trials and worked at the edge of their own inhibitory capacity. We assessed the stop success contrast in accordance with previous IMAGEN publications ^7^.

*Task-based fMRI processing*

Task-based functional MRI data were analyzed with SPM8 (Statistical Parametric Mapping; http://www.fil.ion.ucl.ac.uk/spm). Spatial preprocessing included: slice time correction to adjust for time differences due to multi-slice imaging acquisition, realignment to the first volume in line, non-linearly warping to the MNI space (based on a custom EPI template (53x63x46 voxels) created out of an average of the mean images of 400 adolescents), resampling at a resolution of 3x3x3 mm^3^ and smoothing with an isotropic Gaussian kernel of 5 mm full-width at half-maximum.

*Resting-state fMRI processing*

FEAT (fMRI Expert Analysis Tool) Version 6.00 in FSL was used to perform data preprocessing on the functional data, including motion correction ^22^, slice time correction, BET, spatial smoothing using a Gaussian kernel of 4mm FWHM, and grand mean intensity normalization. Independent Component Analysis (ICA)-based automatic removal of motion-related and physiological noise artifacts was used to further clean the data (ICA-AROMA; ^23^). High-pass temporal filtered (> 0.008 Hz) was applied to  remove slow drifts. The middle EPI volume was co-registered to the individual brain-extracted T1 image using boundary-based registration^24^. Non-linear normalization of the T1 image to the 3mm MNI standard space template (Montreal Neurological Institute, Quebec, Canada) was done using ANTs ^25^. The preprocessed data were normalized to 3mm MNI standard space, applying the registration matrices and warp images from the two previous registration steps. Within-subject time-series were corrected for using Nilearn ([http://nilearn.github.io](http://nilearn.github.io/)) with white matter and cerebrospinal fluid mean activation as covariates.

*Creation of resting-state network matrices*

Resting-state networks were created using FSL’s melodic group independent component analysis (GICA) and network modeling using stage-1 dual regression and the FSLNets toolbox (http://fsl.fMRIb.ox.ac.uk/fsl/fslwiki/FSLNets). Initially, GICA was performed by temporal concatenation of the time-series data from 400 adolescents from IMAGEN to generate 25 independent components. Of those 25 components, 14 were deemed to represent known resting state networks (Supplementary Figure 1). Of note, as little as five dimensionalities are needed to produce similar canonical correlation analysis results as high dimension networks ^26^. The networks were identified using a functional atlas ^27^ and included the: Anterior Salience Network, Auditory Network, Basal Ganglia Network, Dorsal Default Mode Network, Higher Visual Network, Language Network, Left Executive Control Network, Sensorimotor Network, Posterior Salience Network, Precuneus Network, Primary Visual Network, Right Executive Control Network, Ventral Default Mode Network, Visuospatial Network (Supplementary Figure 2). From these 14 networks, FSL’s dual_regression script extracted time-series for each component which was used in FSLNets to calculate a correlation matrix for each subject. In total, there were 91 pairwise correlations (upper-right triangle) that underwent Fischer’s Z-transformation (inverse hyperbolic tangent) to approximately variance-stabilize the correlation coefficients. We did not employ L2 regularization (ridge) because our multiple sparse canonical correlation analysis (see below) already imposes L1 regularization.

*Data driven clustering of task-based fMRI and tract-based spatial statistics*

Given that there is no standard atlas for our fMRI tasks, and the Johns Hopkins University diffusion-based white-matter atlas ^28,29^ does not provide full coverage of our average study tract-based spatial statistics (TBSS) skeleton, we employed hierarchical agglomerative clustering ^30^ using Ward’s algorithm for variance minimization ^31^. We selected this model of hierarchical cluster over other alternatives (e.g., k-means), because for task-based fMRI data Ward’s algorithm performed better than many other geometric clustering methods across a variety of configurations ^32^. Agglomerative clustering was performed using scikit-learn (<https://scikit-learn.org/>). First, to discard low variance noise components, we initially performed a principal component analysis to extract the top 100 components across all subjects, and create a new projected 4D image. Then, using a group data mask, we calculated the adjacency matrix for each voxel in all directions (26 neighbors), and converted it to a sparse matrix in coordinate format. Next, using the de-noised 4D image and the sparse adjacency matrix we applied agglomerative clustering. For the fMRI, we choose 300 clusters within the recommended range of 200-500 (Supplementary Figure 3)^32^. Since we are applying sparsity to the mean values of the clusters, we included whole brain clustering as an additional check to our space canonical correlation model. That is, white matter and cerebrospinal fluid area will be removed from our model. For the TBSS skeleton, we choose 100 clusters since fewer anatomical clusters are likely sufficient for diffusion based signals (Figure S3). Nevertheless, optimizing the number of parcels is still an open question ^32^, especially with respect to TBSS derived metrics.

*Sparse Generalized Canonical Correlation Analysis (SGCCA) data views*

There are a total of eight data views with similar dimensionality (ranging from 91 to 358 variables) including: the clinical scores (335 variables), the EFT task (300 variables), MID task (300 variables), SST task (300 variables), surface area (358 variables), cortical thickness (358 variables), white matter FA (100 variables), and the resting state networks (91 variables). The clinical data were transformed by a negative log transformation (f(x)  = sign(x) * log10(|x| + 1) because the clinical items were left skewed. Since the data views have an unequal number of variables, we weighted each view by dividing by the question of the number of variables. The effect of sex and site was regressed out from all data views using Theil-Sen regression ^33,34^. We used the Theil-sen estimator because it is a non-parametric approach that is robust against potential bias from outliers.

*Estimation of optimal SGCCA hyperparameters*

We used a python wrapper code (<https://github.com/trislett/sparsemodels>) that imports the core RGCCA implementation of the R Penalized Multivariate Analysis (RGCCA) package ^35,36^, and expands on its functionality. For model optimization, we applied a factorial functional scheme (g(x) = x^2^) that maximizes the covariances ^37^. One advantage of the RGCCA package is that it yields orthogonal block (view) components and orthogonal weight vectors (coefficients) for multiview data analyses.

The IMAGEN FU3 sample was divided into training data (70%) and test data (30%) with equal sampling across the eight sites (Figure 1).  Within the training data (n = 599), we optimized the L_1_ sparsity parameter using a permutation scheme for the first component ^38^. We tested ten steps of lambda values ranging from 0.1 to 1.0 in 0.1 steps. For each step, we compared the objective function value of the true model to the rank ordered permuted models to calculate significance, and z-scores for each step were calculated by dividing true values by the standard deviation of the permuted estimates. The best sparsite hyperparameter was selected according to the maximum Z-score. After estimating the optimal sparsity, we then determined the approximate optimal number of components by fitting the SGCCA model with initially 50 components. There is no standard, objective method to selecting the number of components for any matrix factorization methods. Here, we used a similar approach to a principal component analysis, in which we defined the approximate optimal number of components at the point in which the “elbow” of the cumulative average variance explained. After we have estimated the optimal sparsity and number of components, we then applied stability selection. Stability selection is generally applied to sparse CCA since the L_1_ penalties applied to intercorrelated variables can be arbitrary in terms of selection and dependent on prior structure. We choose arguably the most established procedure ^39^ that has previously been applied in the IMAGEN sample ^6^. In brief, a subsample strategy is performed in which half of subjects are randomly selected and a SGCCA model is fit and the variables with non-zero coefficients are saved. The process is repeated 10000 times, and only variables that are selected in 90% of subsampled models are considered stable. Last, a final SGCCA model is fitted to the training data stability selected variables, optimal number of components and without any L1 sparsity.

### Supplementary Table 1. IMAGEN Exclusion Criteria

| Category | Item | Action |
| --- | --- | --- |
| Demographics | Child in target age (14 years) | Inclusion |
|  | Self-reported Western European ancestry | Inclusion |
| Pregnancy and birth | Use of alcohol by the mother during pregnancy (>210 ml alcohol/week) | Exclusion |
|  | Diabetes of the mother during pregnancy (onset before pregnancy, treated  by insulin) | Exclusion |
|  | Premature birth (< 35 weeks) and/or detached placenta | Exclusion |
|  | Hyperbilirubinemia requiring transfusion | Exclusion |
| Child’s medical history | Type 1 diabetes | Exclusion |
|  | Systemic rheumatologic disorders | Exclusion |
|  | Malignant tumors requiring chemotherapy | Exclusion |
|  | Congenital heart defects or heart surgery | Exclusion |
|  | Aneurysm | Exclusion |
| Neurological conditions | Epilepsy | Exclusion |
|  | Bacterial Infection of CNS | Exclusion |
|  | Brain tumor | Exclusion |
|  | Head trauma with loss of consciousness >30 minutes | Exclusion |
|  | Muscular dystrophy, myotonic dystrophy | Exclusion |
| Developmental conditions | Nutritional and metabolic diseases | Exclusion |
|  | Major neuro-developmental disorders (e.g. autism spectrum disorders) | Exclusion |
|  | Hearing deficit | Exclusion |
|  | Vision problems | Exclusion |
| Mental health and abilities | Treatment for schizophrenia, bipolar disorder | Exclusion |
|  | IQ < 70 | Exclusion |
| MR contraindications | Metal implants | Exclusion |
|  | Electronic implants (e.g. pacemakers) | Exclusion |
|  | Severe claustrophobia | Exclusion |

CNS, central nervous system, IQ, Intelligence quotient

### Supplementary Table 2. STRATIFY/ESTRA Exclusion Criteria

| Category | Item | Action |
| --- | --- | --- |
| Demographics | Young adults in target age (18 to 30) | Inclusion |
|  | Self-reported Western European ancestry | Inclusion |
| Medical history | Type 1 or Type 2 diabetes | Exclusion |
|  | Heavily medicated for serious illness (other than diagnosis investigation) | Exclusion |
|  | Participants who are pregnant or any possibility that they may be pregnant | Exclusion |
|  | Restricted mobility, including inability to lie flat for 1.5 hours | Exclusion |
| Neurological conditions | Epilepsy | Exclusion |
|  | Bacterial Infection of CNS | Exclusion |
|  | Brain tumor | Exclusion |
|  | Head trauma with loss of consciousness >30 minutes | Exclusion |
|  | Muscular dystrophy, myotonic dystrophy | Exclusion |
| Developmental conditions | Nutritional and metabolic diseases | Exclusion |
|  | Hearing deficit (requiring hearing aid) | Exclusion |
|  | Vision problems (visual deficit not correctable) | Exclusion |
| MR contraindications | Metal implants | Exclusion |
|  | Electronic implants (e.g. pacemakers) | Exclusion |
|  | Severe claustrophobia | Exclusion |
| Diagnosis Specific Criteria | | |
| Category | Item | Action |
| Healthy Controls | PHQ-9 total score < 5 | Inclusion |
|  | AUDIT total score < 5 | Inclusion |
|  | No current/past mental health issues. | Inclusion |
|  | No regular medication for serious physical health issues. | Inclusion |
|  | No learning difficulties. | Inclusion |
|  | No self-reported regular recreational drug use. | Inclusion |
|  | No 1st or 2nd order family members with mental health issues. | Inclusion |
| Major Depressive Disorder | Current and acute, moderate - severe depression (PHQ-9 >= 15) | Inclusion |
| Alcohol Use Disorder | Moderate - severe alcohol abuse (AUDIT total score >= 15) | Inclusion |
| Psychosis | Schizophrenia, schizotypal and delusional disorders (F20-F29 ICD-10 diagnosis) | Inclusion |
|  | Minimum one-episode schizophreniform illness/ schizophrenia or chronic schizophrenia. | Inclusion |
| Anorexia nervosa | Current diagnosis by EDDS - DSM-5 Version | Inclusion |
|  | BMI < 18.5 (based on self-report of current height and weight). | Inclusion |
| Bulimia nervosa | Current diagnosis by EDDS - DSM-5 Version | Inclusion |

AUDIT, Alcohol Use Disorders Identification Test; BMI, body mass index; CNS, central nervous system; EDDS, Eating Disorder Diagnostic Scale; IQ, Intelligence quotient; PHQ-9, Patient Health Questionnaire-9

### Supplementary Table 3. Sample counts for each data view

| Measure | IMAGEN (BL) | IMAGEN (FU2) | IMAGEN (FU3) | STRATIFY/ESTRA |
| --- | --- | --- | --- | --- |
| DAWBA-SDQ / AUDIT | 2096 | 1300 | 1253 | 485 |
| Emotional Face Task (fMRI) | 1943 | 1403 | 1162 | 511 |
| Stop-Signal Task (fMRI) | 1901 | 1396 | 1157 | 510 |
| Monetary Incentive Delay Task (fMRI) | 1562 | 1339 | 1086 | 347 |
| Cortical Surfaces (CT and SA) | 2012 | 1308 | 1151 | 514 |
| White Matter FA | 1414 | 968 | 942 | 431 |
| Resting-state fMRI | 382 | 1065 | 1168 | 421 |
| Complete Data* | 202 | 683 | 794 | 209 |

AUDIT, Alcohol Use Disorders Identification Test; CT, cortical thickness; DAWBA, Development and Well-Being Assessment; fMRI, functional magnetic resonance imaging, SA, cortical surface area; SDQ, Strength and Difficulties Questionnaire. *The number of subjects with complete data from the clinical data view and all neuroimaging modalities.

### Supplementary Table 4. environMENTAL Consortia members

| Name | Affliation |
| --- | --- |
| Elli Polemiti | Centre of Population Neuroscience and Stratified Medicine (PONS), Department of Psychiatry and Clinical Neuroscience CCM, Charité-Universitätsmedizin Berlin, Berlin, Germany. |
| Esther Hitchen | Centre of Population Neuroscience and Stratified Medicine (PONS), Department of Psychiatry and Clinical Neuroscience CCM, Charité-Universitätsmedizin Berlin, Berlin, Germany. |
| Hedi Kebir | Centre of Population Neuroscience and Stratified Medicine (PONS), Department of Psychiatry and Clinical Neuroscience CCM, Charité-Universitätsmedizin Berlin, Berlin, Germany. |
| Tristram Lett | Centre of Population Neuroscience and Stratified Medicine (PONS), Department of Psychiatry and Clinical Neuroscience CCM, Charité-Universitätsmedizin Berlin, Berlin, Germany. |
| Nilakshi Vaidya | Centre of Population Neuroscience and Stratified Medicine (PONS), Department of Psychiatry and Clinical Neuroscience CCM, Charité-Universitätsmedizin Berlin, Berlin, Germany. |
| Jean-Charles Roy | Centre of Population Neuroscience and Stratified Medicine (PONS), Department of Psychiatry and Clinical Neuroscience CCM, Charité-Universitätsmedizin Berlin, Berlin, Germany. |
| Sören Hese | Institute of Geography, Friedrich Schiller University Jena, Jena, Germany. |
| Paul Renner | Institute of Geography, Friedrich Schiller University Jena, Jena, Germany. |
| Kerstin Schepanski | Institute of Meteorology, Free University Berlin, Berlin, Germany. |
| Jiacan Yuan | Department of Atmospheric and Oceanic Sciences & Institute of Atmospheric Sciences & CMA-FDU Joint Laboratory of Marine Meteorology & IRDR-ICOE on Risk Interconnectivity and Governance on Weather/Climate Extremes Impact and Public Health, Fudan University, Shanghai, China. |
| Gunter Schumann | Centre of Population Neuroscience and Stratified Medicine (PONS), Department of Psychiatry and Clinical Neuroscience CCM, Charité-Universitätsmedizin Berlin, Berlin, Germany; Centre for Population Neuroscience and Stratified Medicine (PONS), Institute for Science and Technology of Brain-inspired Intelligence (ISTBI), Fudan University, Shanghai, China. |
| Tianye Jia | Centre for Population Neuroscience and Stratified Medicine (PONS), Institute for Science and Technology of Brain-inspired Intelligence (ISTBI), Fudan University, Shanghai, China. |
| Xiao Chang | Centre for Population Neuroscience and Stratified Medicine (PONS), Institute for Science and Technology of Brain-inspired Intelligence (ISTBI), Fudan University, Shanghai, China. |
| Yuxiang Dai | Centre for Population Neuroscience and Stratified Medicine (PONS), Institute for Science and Technology of Brain-inspired Intelligence (ISTBI), Fudan University, Shanghai, China. |
| Yunman Xia | Centre for Population Neuroscience and Stratified Medicine (PONS), Institute for Science and Technology of Brain-inspired Intelligence (ISTBI), Fudan University, Shanghai, China. |
| Henrik Walter | Department of Psychiatry and Psychotherapy CCM, Charité-Universitätsmedizin Berlin, Berlin, Germany. |
| Andreas Heinz | Department of Psychiatry and Psychotherapy CCM, Charité-Universitätsmedizin Berlin, Berlin, Germany; German Center for Mental Health (DZPG), Partner Site Berlin-Potsdam, Germany |
| Emin Serin | Department of Psychiatry and Psychotherapy CCM, Charité-Universitätsmedizin Berlin, Berlin, Germany. |
| Markus Ralser | Institute of Biochemistry Charité-Universitätsmedizin Berlin, Berlin, Germany. |
| Sven Twardziok | Berlin Institute of Health at Charité-Universitätsmedizin Berlin, Berlin, Germany. |
| Roland Eils | Berlin Institute of Health at Charité-Universitätsmedizin Berlin, Berlin, Germany. |
| Marcel Jentsch | Berlin Institute of Health at Charité-Universitätsmedizin Berlin, Berlin, Germany. |
| Ulrike Helene Taron | Berlin Institute of Health at Charité-Universitätsmedizin Berlin, Berlin, Germany. |
| Tatjana Schütz | Berlin Institute of Health at Charité-Universitätsmedizin Berlin, Berlin, Germany. |
| Tobias Banaschewski | Department of Child and Adolescent Psychiatry and Psychotherapy, Central Institute of Mental Health, Medical Faculty Mannheim/Heidelberg University, Mannheim, Germany. |
| Maja Neidhart | Department of Psychiatry and Psychotherapy CCM, Charité-Universitätsmedizin Berlin, Berlin, Germany; Department of Child and Adolescent Psychiatry and Psychotherapy, Central Institute of Mental Health, Medical Faculty Mannheim/Heidelberg University, Mannheim, Germany. |
| Nathalie E. Holz | Department of Child and Adolescent Psychiatry and Psychotherapy, Central Institute of Mental Health, Medical Faculty Mannheim/Heidelberg University, Mannheim, Germany. |
| Nina Christmann | Department of Child and Adolescent Psychiatry and Psychotherapy, Central Institute of Mental Health, Medical Faculty Mannheim/Heidelberg University, Mannheim, Germany. |
| Karina Jansone | Department of Child and Adolescent Psychiatry and Psychotherapy, Central Institute of Mental Health, Medical Faculty Mannheim/Heidelberg University, Mannheim, Germany. |
| Andreas Meyer-Lindenberg | Department of Psychiatry and Psychotherapy, Central Institute of Mental Health, Medical Faculty Mannheim/Heidelberg University, Mannheim, Germany. |
| Heike Tost | Department of Psychiatry and Psychotherapy, Central Institute of Mental Health, Medical Faculty Mannheim/Heidelberg University, Mannheim, Germany. |
| Emanuel Schwarz | Department of Psychiatry and Psychotherapy, Central Institute of Mental Health, Medical Faculty Mannheim/Heidelberg University, Mannheim, Germany; Hector Institute for Artificial Intelligence in Psychiatry, Central Institute of Mental Health, Medical Faculty Mannheim/Heidelberg University, Mannheim, Germany. |
| Frauke Nees | Institute of Medical Psychology and Medical Sociology, University Medical Center Schleswig Holstein, Kiel University, Kiel, Germany. |
| Sebastian Siehl | Institute of Medical Psychology and Medical Sociology, University Medical Center Schleswig Holstein, Kiel University, Kiel, Germany. |
| Stephan Lehmler | Institute of Medical Psychology and Medical Sociology, University Medical Center Schleswig Holstein, Kiel University, Kiel, Germany. |
| Ole A. Andreassen | Centre for Precision Psychiatry, Division of Mental Health and Addiction, Oslo University Hospital & Institute of Clinical Medicine, University of Oslo, Oslo, Norway. |
| Lars T. Westlye | Centre for Precision Psychiatry, Division of Mental Health and Addiction, Oslo University Hospital & Institute of Clinical Medicine, University of Oslo, Oslo, Norway; Department of Psychology, University of Oslo, Oslo, Norway. |
| Dennis van der Meer | Centre for Precision Psychiatry, Division of Mental Health and Addiction, Oslo University Hospital & Institute of Clinical Medicine, University of Oslo, Oslo, Norway;School of Mental Health and Neuroscience, Faculty of Health, Medicine and Life Sciences, Maastricht University, The Netherlands. |
| Sara Fernández-Cabello | Centre for Precision Psychiatry, Division of Mental Health and Addiction, Oslo University Hospital & Institute of Clinical Medicine, University of Oslo, Oslo, Norway; Department of Psychology, University of Oslo, Oslo, Norway. |
| Rikka Kjelkenes | Centre for Precision Psychiatry, Division of Mental Health and Addiction, Oslo University Hospital & Institute of Clinical Medicine, University of Oslo, Oslo, Norway. |
| Michael Rapp | Department for Social and Preventive Medicine, University of Potsdam, Potsdam, Germany. |
| Mira Tschorn | Department for Social and Preventive Medicine, University of Potsdam, Potsdam, Germany. |
| Sarah J. Böttger | Department for Social and Preventive Medicine, University of Potsdam, Potsdam, Germany. |
| Andre Marquand | Donders Institute/Radboud UMC, Nijmegen, the Netherlands. |
| Antoine Bernas | Donders Institute/Radboud UMC, Nijmegen, the Netherlands. |
| Gaia Novarino | Institute of Science and Technology Austria (ISTA), Klosterneuburg, Austria. |
| Mel Slater | Campus de Mundet, ICREA-University of Barcelona, Barcelona, Spain; Department of Computer Science, University College London, London, United Kingdom. |
| Jaime Gallego | Campus de Mundet, ICREA-University of Barcelona, Barcelona, Spain. |
| Álvaro Pastor | Campus de Mundet, ICREA-University of Barcelona, Barcelona, Spain. |
| Peter Sommer | Ksilink, Strasbourg, France. |
| Karen Schmitt | Ksilink, Strasbourg, France. |
| Johannes H. Wilbertz | Ksilink, Strasbourg, France. |
| Viktor Jirsa | Institut National de la Santé et de la Recherche Médicale (Inserm), Institut de Neurosciences des Systèmes (INS) UMR1106, Aix Marseille Université, Marseille, France. |
| Spase Petkoski | Institut National de la Santé et de la Recherche Médicale (Inserm), Institut de Neurosciences des Systèmes (INS) UMR1106, Aix Marseille Université, Marseille, France. |
| Anastasios-Polykarpos Athanasiadis | Institut National de la Santé et de la Recherche Médicale (Inserm), Institut de Neurosciences des Systèmes (INS) UMR1106, Aix Marseille Université, Marseille, France. |
| Vince D. Calhoun | Tri-institutional Center for Translational Research in Neuroimaging and Data Science (TReNDS), Georgia State, Georgia Tech, Emory, Atlanta, Georgia. |
| Nicholas Clinton | Google, Inc, Mountain View, California. |
| Sylvane Desrivières | Institute of Psychiatry, Psychology & Neuroscience, SGDP Centre, King's College London, London, United Kingdom. |
| Kofoworola Agunbiade | Institute of Psychiatry, Psychology & Neuroscience, SGDP Centre, King's College London, London, United Kingdom. |
| Xinyang Yu | Institute of Psychiatry, Psychology & Neuroscience, SGDP Centre, King's College London, London, United Kingdom. |
| Zuo Zhang | Institute of Psychiatry, Psychology & Neuroscience, SGDP Centre, King's College London, London, United Kingdom. |
| Di Chen | Institute of Psychiatry, Psychology & Neuroscience, SGDP Centre, King's College London, London, United Kingdom. |
| Georgie Keggin | Institute of Psychiatry, Psychology & Neuroscience, SGDP Centre, King's College London, London, United Kingdom. |
| Ameli Schwalber | Concentris research management GmbH. |
| Dennis Cleff | Concentris research management GmbH. |
| Bernd Carsten Stahl | School of Computer Science, University of Nottingham, Nottingham, United Kingdom. |
| George Ogoh | School of Computer Science, University of Nottingham, Nottingham, United Kingdom. |
| Tamara Schikowski | IUF – Leibniz Research Institute for Environmental Medicine, Düsseldorf, Germany. |
| Ragnhild Eek Brandlistuen | Norwegian Institute of Public Health, Oslo, Norway. |
| Guillem Feixas | Department of Clinical Psychology and Psychobiology, Universitat de Barcelona, Barcelona, Spain; Institute of Neurosciences, Universitat de Barcelona, Barcelona, Spain. |
| Francisco J. Eiroa-Orosa | Department of Clinical Psychology and Psychobiology, Universitat de Barcelona, Barcelona, Spain; Institute of Neurosciences, Universitat de Barcelona, Barcelona, Spain. |
| Per Hoffmann | Institute of Human Genetics, University Hospital of Bonn, Bonn, Germany. |
| Markus M. Nöthen | Institute of Human Genetics, University Hospital of Bonn, Bonn, Germany. |
| Andreas J. Forstner | Institute of Human Genetics, University Hospital of Bonn, Bonn, Germany. |
| Abigail Miller | Institute of Human Genetics, University Hospital of Bonn, Bonn, Germany. |
| Carina M. Mathey | Institute of Human Genetics, University Hospital of Bonn, Bonn, Germany. |
| Isabelle Claus | Institute of Human Genetics, University Hospital of Bonn, Bonn, Germany. |
| Stefanie Heilmann-Heimbach | Institute of Human Genetics, University Hospital of Bonn, Bonn, Germany. |
| Yuzhu Li | Institute of Science and Technology for Brain-Inspired Intelligence, Fudan University, Shanghai, China. |
| Yanqing Zhang | Institutes of Biomedical Sciences, Fudan University, Shanghai, China. |
| Argyris Stringaris | Divisions of Psychiatry and Division of Clinical, Educational & Health Psychology, University College London, London, UK; First Department of Psychiatry, Aiginiteion Hospital, National and Kapodistrian University of Athens. |

### Supplementary Figure 1. Ward’s hierarchical clustering of the 14 resting state network edges and correlation matrix of the dual regression synthetic time series.

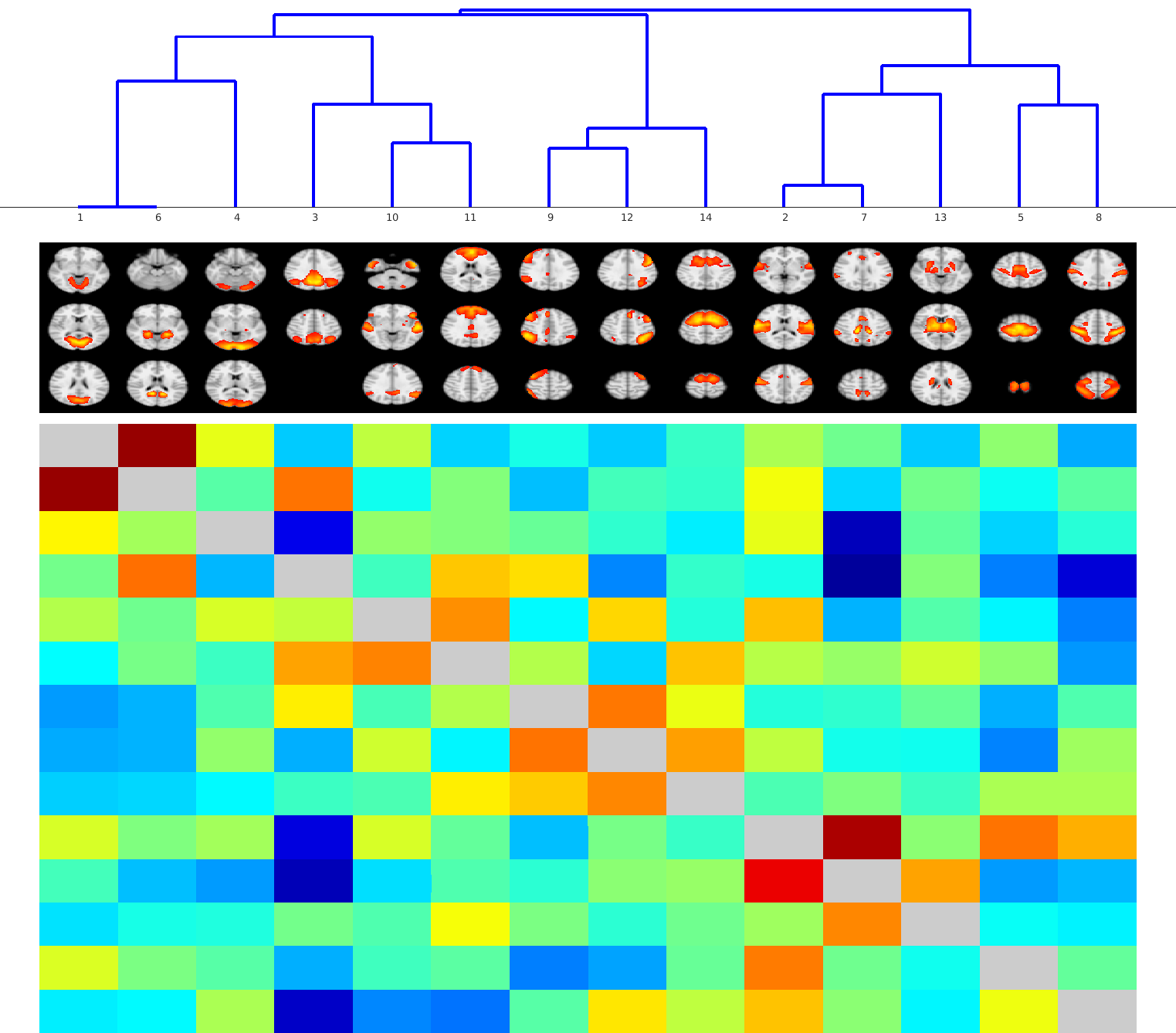

Identified resting state networks are numbered as the following: Anterior Salience Network (14), Auditory Network (2), Basal Ganglia Network (13), Dorsal Default Mode Network (11), Higher Visual Network (4), Language Network (10), Left Executive Control Network (12), Sensorimotor Network (5), Posterior Salience Network (7), Precuneus Network (3), Primary Visual Network (1), Right Executive Control Network (9), Ventral Default Mode Network (6), Visuospatial Network (8).

### Supplementary Figure 2. The parcels derived from Ward’s hierarchical clustering

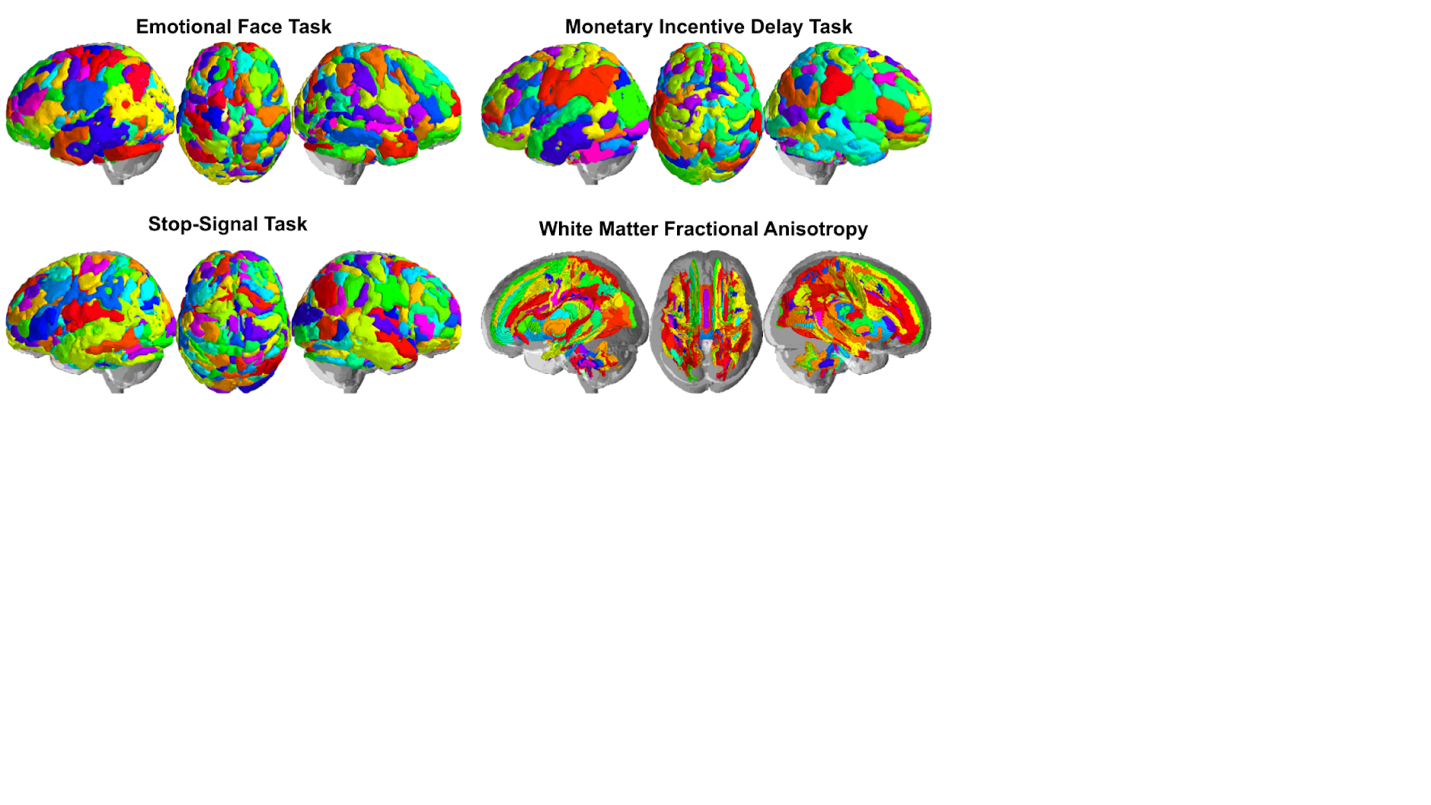

Ward’s hierarchical clustering label for the emotional face fMRI task, monetary incentive delay fMRI, stop-signal fMRI task, and white matter fractional anisotropy TBSS skeleton. Each region of the atlases is assigned random colors.

### Supplementary Figure 3. Longitudinal association between psychopathology scores and neuroimaging scores in training and test data.

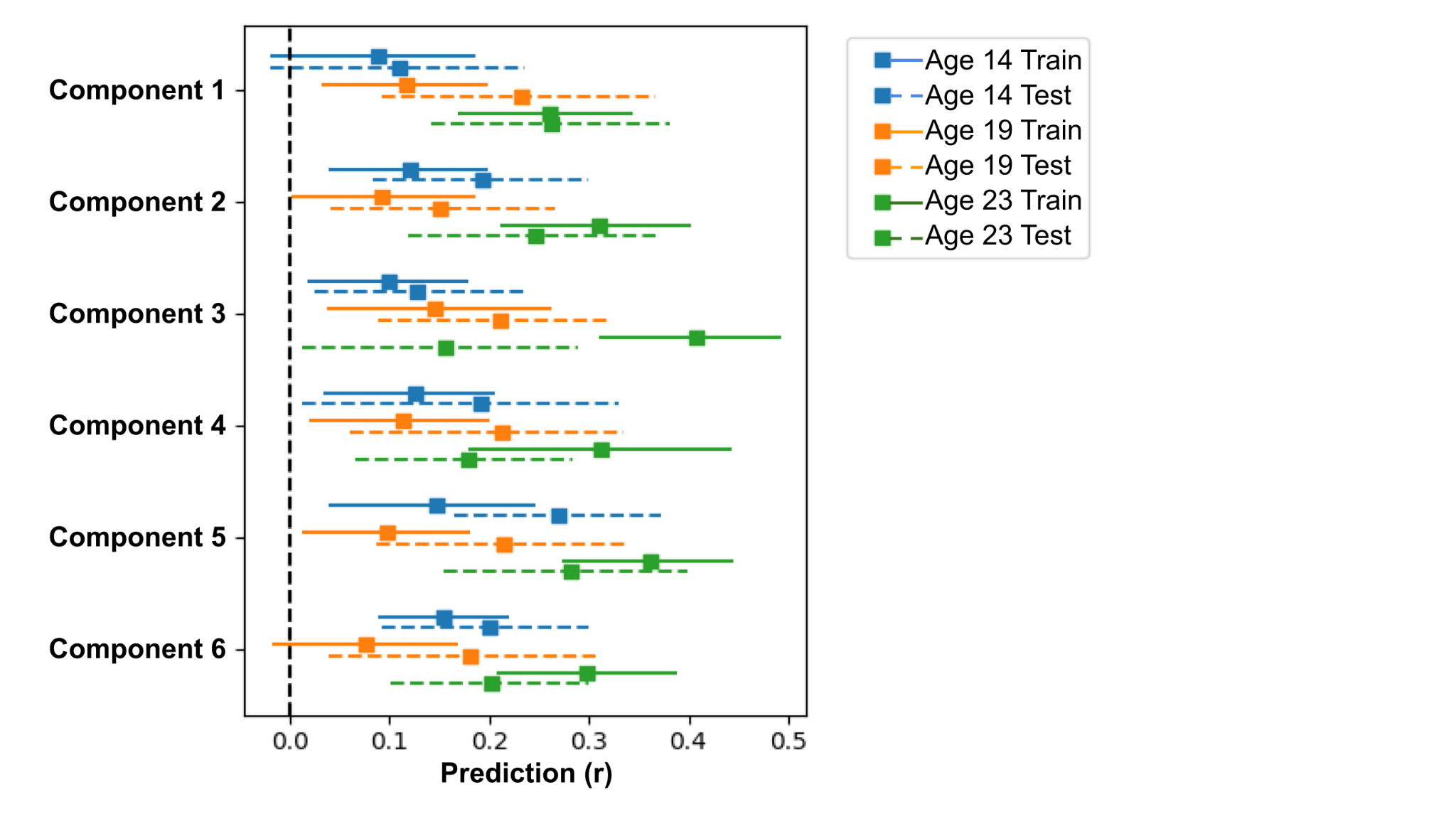

To evaluate the longitudinal relationship between clinical symptoms and neuroimaging features, SGCCA-regression was employed. The endogenous variables were the clinical view scores, and the neuroimaging view scores were the exogenous variables corresponding to each component. The figure includes bootstrapped 95% confidence intervals represented by blue, yellow, and red lines at ages 14, 19, and 23. Solid lines represent training data, while the dotted lines represent test data. The solid box indicates the canonical correlation value (r). Components 1 to 6 correspond to components with manic, depressive, anxiety, stress, eating disorder, and panic attack clinical symptoms. Notably, the green lines correspond to the same results as in Figure 4.
